# Genetic ancestry superpopulations show distinct prevalence and outcomes across pediatric central nervous system tumors from the PBTA and PNOC

**DOI:** 10.1101/2024.06.14.24308885

**Authors:** Ryan J. Corbett, Cricket C. Gullickson, Zhuangzhuang Geng, Miguel A. Brown, Bo Zhang, Chuwei Zhong, Nicholas Van Kuren, Antonia Chroni, Christopher Blackden, Ammar S. Naqvi, Alexa Plisiewicz, Sean McHugh, Emmett Drake, Kaitlin Lehmann, Tom B. Davidson, Michael Prados, Phillip B. Storm, Adam C. Resnick, Angela J. Waanders, Sebastian M. Waszak, Sabine Mueller, Jo Lynne Rokita, Cassie Kline

## Abstract

**Background:** Central nervous system (CNS) tumors lead to cancer-related mortality in children. Genetic ancestry-associated cancer prevalence and outcomes have been studied, but is limited.

**Methods:** We performed genetic ancestry prediction in 1,454 pediatric patients with paired normal and tumor whole genome sequencing from the Open Pediatric Cancer (OpenPedCan) project to evaluate the influence of reported race and ethnicity and ancestry-based genetic superpopulations on tumor histology, molecular subtype, survival, and treatment.

**Results:** Predicted superpopulations included African (AFR, N=153), Admixed American (AMR, N=223), East Asian (EAS, N=67), European (EUR, N=968), and South Asian (SAS, N=43). Reported race and ethnicity and ancestry-based genetic superpopulations were non-randomly associated. Patients with an atypical teratoid rhabdoid tumor or meningioma were enriched for AFR ancestry. Patients of AMR ancestry with *KIAA1549::BRAF* fusion-positive low-grade glioma (LGG) had tumors enriched for rare fusion breakpoints, lesser extent of surgical resection, and worse event-free survival (EFS). Non-EUR and AMR patients with germ cell tumors or SHH-activated medulloblastoma exhibited worse EFS relative to EUR patients, and patients of AFR ancestry with LGG or ependymoma had worse overall survival compared to EUR patients. We observed higher frequency of clinical trial enrollment among AMR patients across tumor histologies, but increased utilization of photon versus proton radiation relative to other superpopulations.

**Conclusions:** Genetic ancestry-associated differences exist across pediatric CNS tumor histological and molecular subtypes. Further investigation into genetic and socioeconomic factors contributing to these observed inequities is needed.

**Key Points:** Distinct associations of genetic ancestry-based superpopulations exist within pediatric CNS tumor histologic and molecular subtypes and correlate with survival outcomes and treatment.

**Importance of the Study:** This work provides critical insight on the impact of reported race and ethnicity and genetic-based ancestry superpopulations on nearly 1,500 pediatric patients with CNS tumors who had matched normal and tumor sequencing performed. We identify novel associations between ancestry superpopulations and tumor histology, molecular subtypes, and treatments received. Here, we begin to inform on the contributions of social constructs of race and ethnicity and tumor characteristics that are enriched among genetic-based ancestry superpopulations on clinical outcomes of pediatric patients with CNS tumors. Our findings indicate that potential social and genetic risk stratifications exist for pediatric CNS tumors and warrant further investigation to ensure equitable clinical outcomes for all patients.

## Introduction

Primary central nervous system (CNS) tumors are the leading cause of cancer-related mortality in children^1,2^. Previous work has shown that overall and tumor-specific survival outcomes and incidence vary according to race and ethnicity in children with primary CNS tumors^1,2^. For example, while White children have higher incidences of CNS tumors in general, they have lower incidences of malignant CNS tumors compared to other races^1,3^. Furthermore, Black and Hispanic patients have higher rates of mortality compared to White patients, and Hispanic children are more likely to present with advanced diseases^1,2,4^. Prior studies have attempted to characterize the contribution of sociodemographic factors, such as socioeconomic status, on survival outcomes and treatment strategies while accounting for key variables like extent of disease, type of treatment, and age at diagnosis. Even in consideration of these confounding variables, findings demonstrate differences in survival and treatment according to patient race and ethnicity^3,5,6^. This suggests that unmeasured social determinants of health (SDoH) or inherent genetic variation in cancer risk may be playing a role^3^. What remains less well understood is the individual contribution of these distinct outcomes as it relates to environmental exposures, pediatric CNS tumor type, molecular subtype, and clinical characteristics.

It is essential to recognize that race and ethnicity are social and cultural constructs distinct from genetic ancestry, which can be estimated using genetic markers that capture ancestral population migration patterns and admixture events^7^. The use of predicted genetic ancestry in cancer studies has revealed numerous ancestry-based correlates to cancer incidence and outcome. Work using data from The Cancer Genome Atlas (TCGA) showed increased frequency of *TP53* mutations in patients of African ancestry with cancer types demonstrating high chromosomal instability, as well as decreased frequency of *VHL* and *PBRM1* mutations in renal cancer patients of African ancestry^8,9^. In a study of pediatric acute lymphoblastic leukemia (ALL), East Asian ancestry was negatively associated with *BCR-ABL1*-like and T-cell ALL incidences, while increased proportion of African and Native American genetic ancestry was associated with worse overall and event-free survival (OS and EFS, respectively)^10^. Further, within pediatric brain tumors, prior reports have demonstrated linkages between ancestry, self-identified ethnicity, and potential genomic loci of germline risk in pilocytic astrocytoma and ependymoma^11,12^. While concordance between reported race and ethnicity and predicted genetic ancestry can vary by group, prior work has reported a significant non-random association between these categories^13^. Thus, the use of genetic ancestry to assess demographic inequities in cancer outcomes may be particularly useful in cohorts for which electronic medical records are incomplete or inaccurate^7^.

In the current study, we aim to go beyond previous work investigating genomic correlates of cancer risk in isolated silos and utilize a large cohort of patients with broad histologies of primary pediatric CNS tumors to explore the potential contributions of genomics and social health risk categories. We specifically utilize predicted genetic ancestries to evaluate associations with incidence and clinical outcomes across a diverse group of pediatric CNS tumor diagnoses. This work provides an essential framework to better characterize the contributions of genetic and sociodemographic factors to cancer outcomes in patients with pediatric CNS tumors, and ideally augment our understanding of pediatric CNS tumor risk stratification through a lens of health equity.

## Materials & Methods

### Pediatric CNS tumor patient cohort

The pediatric CNS tumor patient cohort used in this study was derived from the Open Pediatric Cancer (OpenPedCan) project^14,15^, an open analysis effort at the Children’s Hospital of Philadelphia that performs pediatric cancer data harmonization and shares results from downstream analyses. Pediatric Brain Tumor Atlas (PBTA) and Pediatric Neuro-Oncology Consortium (PNOC) patients under 40 years of age with matched tumor and normal whole genome sequencing (WGS) from OpenPedCan at the time of data release v12 were included for this study. These included patients from the Children’s Brain Tumor Network (CBTN, https://cbtn.org/<u>, N=1,356</u>), PNOC (https://pnoc.us/, N=35), Oligo Nation (https://www.oligonation.org/, N=30), and Project HOPE (an adolescent and young adult high-grade glioma study, N=33). A full list of participating institutions from which these samples were collected can be found in **Table S1**. Patients from countries outside the United States were excluded due to consent procedures necessary to ascertain race and ethnicity data as per local regulatory requirements. We pulled demographics and clinical data including reported race and ethnicity, initial tumor diagnosis, tumor histology, molecular subtype, survival outcomes (EFS and OS), and selected treatment data from OpenPedCan data release v14. A detailed overview of molecular subtyping methods can be found in Shapiro et al. 2023^14^ with updates to high-grade glioma (HGG) and atypical teratoid rhabdoid tumor (ATRT) described in OpenPedCan^15^. Collected treatment information through the IRB-approved CBTN project included extent of tumor resection, utilization of upfront proton versus photon therapy for those who received radiation therapy, and clinical trial enrollment at time of diagnosis. Patient data from the PNOC cohort, such as demographics, diagnostic data, molecular sequencing, and survival, was collected within confines of clinical trial data collection and shared as per IRB-approved consent. Reported race and ethnicity were utilized as entered by local site investigators and research teams.

### Predicted ancestry

We used the somalier suite of tools (v.0.2.15) to predict ancestry superpopulations from non-tumor WGS data^16^. First, ‘somalier-extract‘ was applied to alignment CRAM files to obtain variant calls at known polymorphic sites for each sample. We then used ‘somalier-ancestry‘ with default parameters to estimate ancestry superpopulations in patients using genetic markers from reference individuals of known ancestry from the 1000 Genomes Project^17^. Briefly, dimensionality reduction was performed on query and reference genotype data to estimate five principal components (PCs). The resulting PC values were used to estimate the proportion of genetic ancestry assigned to each of five ancestry superpopulations defined by the 1000 Genomes Project: Sub-Saharan African (AFR), Admixed American (AMR), East Asian (EAS), European (EUR), and South Asian (SAS). We use the term “Admixed American” here to emphasize the known European, African, and Native-American admixture that is observed in individuals sampled from populations in the Americas by the 1000 Genomes Project^17^. The ancestry group with the highest estimated percentage in each sample was assigned as the predicted genetic ancestry superpopulation.

### BRAF fusion breakpoint analyses for low-grade glioma

All high-confidence, in-frame *KIAA1549*::*BRAF* STAR-fusion and/or Arriba fusion calls in pediatric low-grade glioma (LGG) tumors were annotated with exon number in canonical transcripts (NM_001164665 for *KIAA1549* and NM_004333 for *BRAF*) using *biomaRt* and *GenomicRanges* R packages^18–20^. Common breakpoints included those involving exons 15:09 (*KIAA1549* exon 15 and *BRAF* exon 9), 16:09, 16:11, and 18:10^21,22^. All other breakpoint combinations were classified as rare.

### Statistical analyses

The association between reported race and ethnicity and genetic ancestry superpopulation was assessed using Fisher’s exact tests. Enrichment of superpopulations within race and ethnicity groups was calculated using hypergeometric tests. We integrated CNS tumor histology and molecular subtype data from matched tumor samples with survival data to determine whether genetic ancestry superpopulations were associated with cancer type, subtype, and/or survival. Non-random distribution of superpopulations within histologies and molecular subtypes was assessed using Fisher’s exact tests and enrichment was calculated using hypergeometric tests. Statistical significance was defined as Benjamini-Hochberg-adjusted FDR<0.05.

We performed Kaplan-Meier analysis of OS and EFS within histologies and molecular subtypes to compare outcomes of patients of different superpopulations. We generated Cox proportional-hazards regression models to identify additional variables that were predictive of survival using the following covariates: age at diagnosis (all models), molecular subtype (ATRT, LGG, HGG, ependymoma [EPN], medulloblastoma [MB], mixed glioneuronal tumors), and extent of tumor resection (ATRT, LGG, EPN, HGG, MB, and schwannoma, to accommodate known impact of degree of resection on outcome in these cohorts). Analyses of deviance were performed on Cox proportional-hazards models to assess overall effect of genetic ancestry superpopulation on survival. All survival analyses were performed using the *survival* R package^23^, and Kaplan-Meier survival curves were generated using the *survminer* R package^24^.

## Results

### Ancestry superpopulation prediction summary

We predicted genetic ancestry for 1,454 pediatric CNS tumor patients from the PBTA and PNOC. Patients were classified into genetic ancestry superpopulations as follows: N=153 AFR, N=223 AMR, N=67 EAS, N=968 EUR, and N=43 SAS (**Figure 1A, Table S2**). Most patients (1276/1454; 87.8%) exhibited superpopulation probabilities greater than 90% (**Figure S1A**). The remaining 178 patients all displayed a non-zero probability of AMR ancestry, with 51 patients classified into the AMR superpopulation and 127 classified into a non-AMR superpopulation (EUR=91, AFR=32, EAS=3, SAS=1; **Figure S1B**).

**Figure 1.**
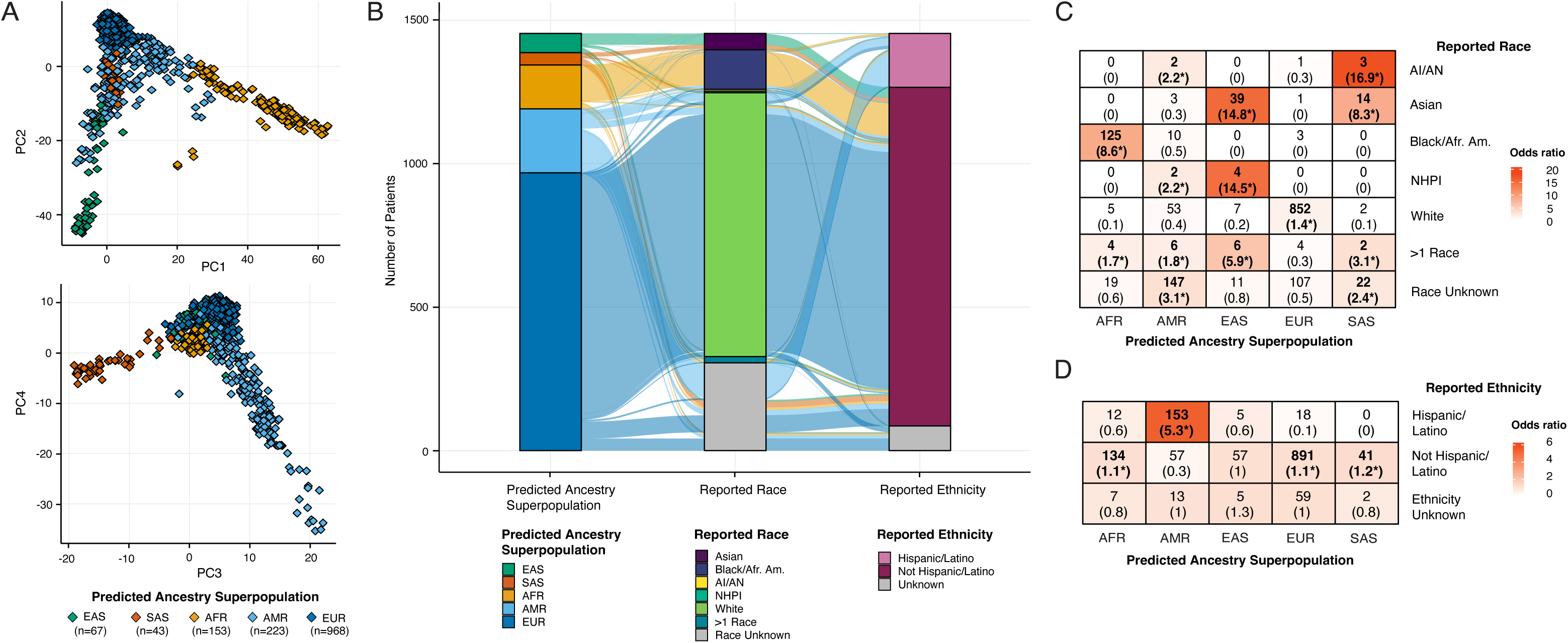
Genetic ancestry prediction in a pediatric central nervous system tumor cohort. **A.** Somalier genetic ancestry prediction principal component (PC) plot of PCs 1-4. **B.** Alluvial plot displaying relationships between genetic ancestry superpopulation, reported race and ethnicity in pediatric CNS tumor cohort. **C-D**. Count of genetic ancestry superpopulation members within each (**C**) reported race and (**D**) ethnicity category. Fisher’s exact test-derived odds ratios are noted in parentheses with stars denoting Fisher’s exact test FDR<0.05. Cells are colored by odds ratio weight.

There was a significant non-random association between reported race and ethnicity and superpopulation (p<0.001; **Figure 1B, Table S3**). Patients of each superpopulation were significantly enriched for distinct reported race groups (**Figure 1C**). This included AFR patients for Black/African American race, EAS patients for Asian and NHPI races, EUR patients for White race, and SAS patients for AI/AN and Asian races. AMR patients were significantly enriched among patients of reported Hispanic/Latino ethnicity, while non-AMR patients were enriched among patients of reported non-Hispanic/Latino ethnicity (**Figure 1D**). We predicted genetic ancestry superpopulations in 328 individuals (22.5% of participants) for which reported race or ethnicity data were unavailable, and these patients were disproportionately assigned to AMR and SAS superpopulations (AMR: N=147, OR=3.1, FDR=4.9e-58; SAS: N=22, OR=2.4, FDR=7.4e-06).

### Genetic ancestry superpopulations are enriched for distinct pediatric CNS tumor histologies

We identified a significant non-random association between genetic ancestry superpopulation and CNS tumor histology (p=1.0e-04; **Figure 2A, Table S3**). Patients with ATRT and meningiomas were significantly enriched within the AFR superpopulation (N=11, OR=2.2, FDR=0.01; N=10, OR=2.3, FDR=0.01, respectively). We observed a significant enrichment of patients with DIPG or DMG within the AMR superpopulation (N=28, OR=1.7, FDR=0.01). To determine if this enrichment was due to inclusion of patients enrolled in PNOC trials—of which the majority (34/35, 97.1%) had DIPG or DMG—we assessed superpopulation distribution among these patients. The AMR superpopulation was significantly enriched among PNOC patients (N=14, OR=3.9, p=2.6e-04). Furthermore, re-analysis of tumor histology enrichment among superpopulations when excluding PNOC patients resulted in loss of AMR superpopulation enrichment in patients with DIPG or DMG (OR=1.3, FDR=0.5), indicative of a patient sampling bias. Patients with germ cell tumors (GCTs) were significantly enriched among the EAS superpopulation (N=4, OR=3.8, FDR=0.02), with three patients diagnosed with teratomas and one with a mixed GCT. Patients with LGG and mixed neuronal-glial tumors were significantly enriched within the EUR superpopulation (N=275, OR=1.1, FDR=0.02; N=93, OR=1.1, FDR=0.04, respectively). Lastly, patients with schwannomas were significantly enriched among SAS patients (N=4, OR=5.2, FDR=3.7e-03).

**Figure 2.**
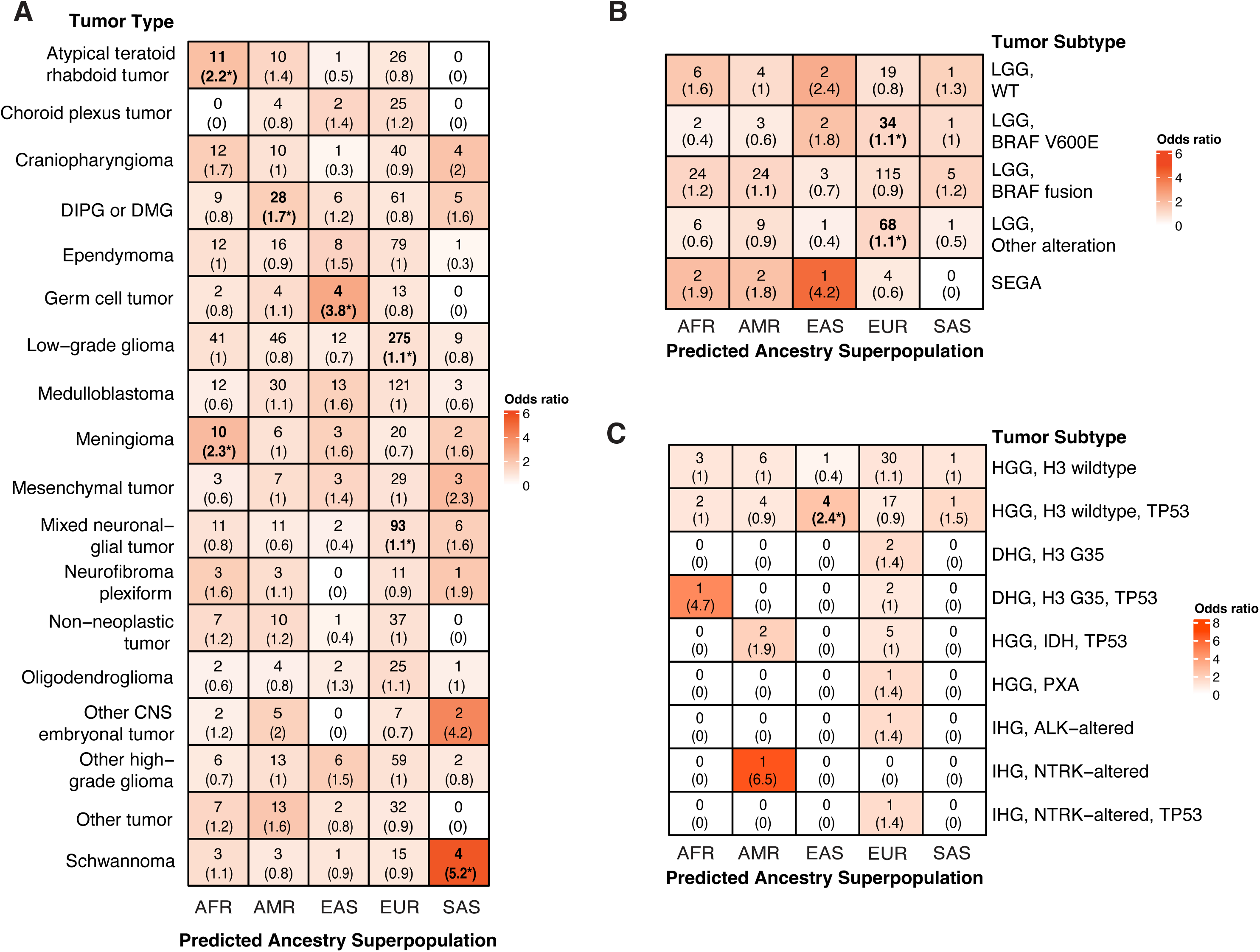
Distribution of CNS tumor histologies and molecular subtypes across genetic ancestry superpopulations. **A.** Count of predicted genetic ancestry superpopulation members within CNS tumor histology cohorts, with Fisher’s exact test-derived odds ratios in parentheses. Cells are colored by odds ratio weight. Stars denote FDR<0.05. **B-C.** Count and enrichment of predicted ancestry superpopulation members within low-grade glioma (LGG, B) and (HGG, C) molecular subtype cohorts. Stars denote FDR<0.05. DHG = diffuse hemispheric glioma, PXA = pleiomorphic xanthoastrocytoma, IHG = infant-type hemispheric glioma.

### Novel molecular findings can be found across genetic ancestry superpopulations

We observed significant enrichment of genetic ancestry superpopulations in molecular subtypes of specific CNS tumor histologies. We found that patients with non-*BRAF* altered LGG (“Other alteration”) tumors were enriched among EUR patients (N=34, OR=1.1, FDR=0.047); **Figure 2B**). Furthermore, EAS patients with HGG exhibited higher relative incidence of H3 wildtype, *TP53*-altered tumors (N=4, OR=2.4, FDR=0.01). We did not observe superpopulation-specific enrichment of molecular subtypes within other tumor histologies (**Figure S2**).

A recent report from the PNOC001 trial identified two classes of in-frame *KIAA1549*::*BRAF* fusion breakpoints in LGG tumors denoted as “common” (16:09, 15:09, 16:11, and 18:10) or “rare” (any other combination), with rare breakpoints associated with supratentorial midline pilocytic astrocytoma (PA) and poor clinical outcomes^22^. Since LGGs made up the largest histology of this cohort and we had a large number of patients with *KIAA1549*::*BRAF* fusions (N=172/383), we sought to determine whether ancestral associations with breakpoint type exist. All *KIAA1549::BRAF* fusion-positive LGGs were confirmed to have a PA (N=162), pilomyxoid astrocytoma (PMA, N=8), or fibrillary astrocytoma (N=2) diagnosis. The majority of tumors (161/172, 93.6%) harbored one of the four “common” breakpoints, ranging from 87.5% (21/24) of AMR patients to 100% of EAS and SAS patients. Prevalence of the 15:09 breakpoint was not evenly distributed across superpopulations (p=0.04), with EAS and SAS patients being significantly enriched for 15:09 breakpoints relative to other superpopulations (EAS: N=2, OR=3.7, FDR=0.01; SAS: N=3, OR=3.3, FDR=0.01; **Figure 3A**). AMR patients disproportionately harbored rare fusion breakpoints, but this was not significant after correcting for multiple tests (N=3, OR=2.0, FDR=0.25).

**Figure 3.**
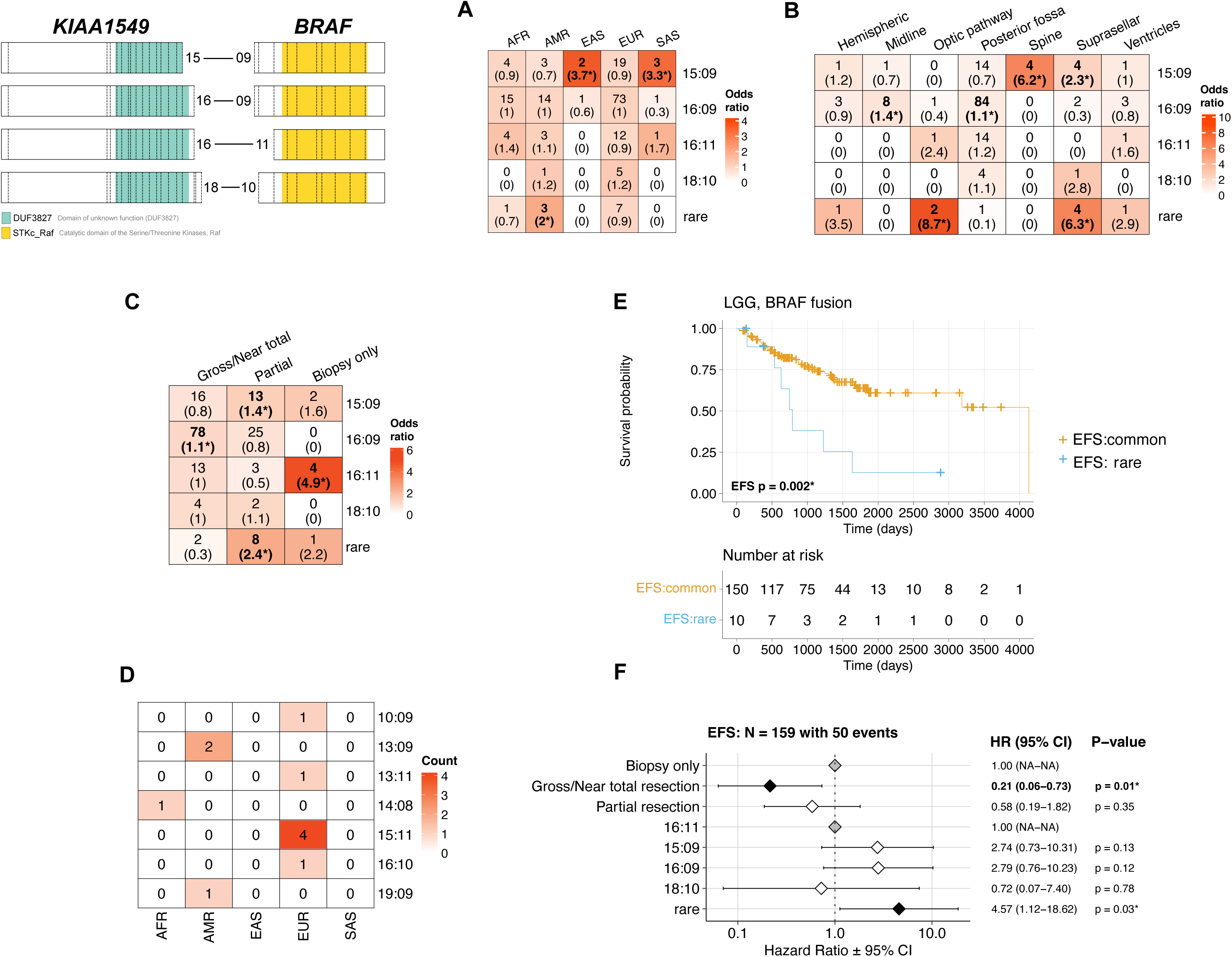
*KIAA1549*::*BRAF* fusion breakpoint distribution by genetic ancestry. A-C. Count of *KIAA1549*::*BRAF* fusion breakpoint group tumors by (**A**) genetic ancestry superpopulation, (**B**) CNS anatomic location, and (**C**) degree of tumor resection, with Fisher’s exact test-derived odds ratios in parentheses. Cells are colored by odds ratio weight. Stars denote FDR<0.05. **D.** Distribution of rare and novel fusion breakpoints by genetic ancestry superpopulation. **E.** Kaplan-Meier EFS curves for patients with LGG *BRAF* fusions harboring common or rare breakpoints. **F.** Cox proportional hazards model forest plots of EFS in LGG *BRAF* fusion cohort, including covariates for tumor resection level and breakpoint type (common vs. rare). Grey diamonds represent the reference (control) groups, black diamonds represent a significant difference between a group and the reference (p < 0.05), and white diamonds represent non- significant differences between a group and the reference. Listed for each group are the hazard ratio (HR) with 95% confidence intervals and p-values for each term. A p-value < 0.05 for a given group level in forest plots indicate a significant difference in relative risk of an event relative to that of the reference level of group.

We also observed breakpoint type-specific distribution of LGG tumors in CNS regions that correlated with extent of tumor resection (**Figure 3B-C**). Tumors with *KIAA1549::BRAF* 16:09 breakpoints were significantly enriched in the posterior fossa (N=84, OR=1.1, FDR=3.0e-03), and these were significantly more likely to undergo total resection than other breakpoint types (N=78, OR=1.1, FDR=8.9e-04). Conversely, tumors with 15:09 breakpoints were enriched in suprasellar and spinal regions (N=4, OR=2.3, FDR=0.06; N=4, OR=6.2, FDR=5.3e-03), and those with rare breakpoints were significantly enriched in suprasellar and optic pathway regions (N=4, OR=6.3, FDR=4.9e-04; N=2, OR=8.7, FDR=1.8e-03). Both 15:09 and rare

*KIAA1549::BRAF* breakpoint tumors were more likely to undergo partial resection relative to tumors with other breakpoint types (15:09: N=13, OR=1.4, FDR=0.1; rare: N=8, OR=2.4, FDR=1.1e-03). Lastly, 15:09 and rare breakpoint tumors were significantly more likely to be diagnosed as PMAs versus PAs (15:09: N=4, OR=4, FDR=0.03; rare: N=3, OR=5.2, FDR=5.0e- 03), whereas other common breakpoint tumors were more likely to be diagnosed as PAs (16:09: N=102, OR=1.1, FDR=6.4e-05; **Figure S3**)

We identified eleven patients with seven distinct rare *KIAA1549::BRAF* breakpoints, two of which were recurrent and unique to a single superpopulation: 13:09 breakpoints were identified in tumors from two AMR patients, and 15:11 breakpoints were identified in tumors from four EUR patients (**Figure 3D**). To determine if breakpoint type was associated with survival differences, we generated survival models for EFS using breakpoint type as a predictor. Significantly worse EFS persisted in patients with rare *KIAA1549::BRAF* breakpoints relative to those with 16:11 breakpoints, even when accounting for degree of tumor resection (1.9 versus 2.9 years, HR=5.3, p=0.03, **Figure 3E-F**).

### Patients from non-European genetic ancestry superpopulations have significantly worse event-free and overall survival in a subset of pediatric CNS tumors

We calculated median EFS and OS for each CNS tumor histology and molecular subtype with sufficient sample size (N≥20) and number of recorded events (N≥10) across genetic ancestry superpopulations (**Table S4**). A summary of analyses of deviance on cox proportional hazards models is shown in **Table S4**. A liberal p-value (<0.1) was applied to select a subset of tumor histologies to further explore superpopulation-specific survival differences. We observed correlations between superpopulation and EFS in GCT (χ=12.4, p=0.01), neurofibroma plexiform (χ=7.6, p=0.02), schwannoma (χ=11.4, p=0.02), and MB, SHH-activated and Group 3 (χ=6.6, p=0.09; χ=7.1, p=0.07) cohorts. Furthermore, OS was correlated with superpopulation in EPN (χ=4.6, p=0.09), LGG (χ=11.4, p=0.02), and mesenchymal tumor (χ=14.0, p=0.01) cohorts. Next, we assessed pairwise differences in patient survival by superpopulation within the cohorts selected above. Among patients with GCTs, those from non-EUR superpopulations exhibited significantly worse EFS relative to those from the EUR superpopulation (median EFS 0.5 vs. 2.1 years, HR=12, p=0.01; **Figure 4A-B**). Patients with MB-SHH subtype tumors from the AMR superpopulation exhibited significantly worse EFS relative to EUR patients (median EFS 1.0 vs. 2.9 years, HR=4.3, p=0.04; **Figure 4C-D**). We did not find evidence of non-random distribution of the four MB SHH subtypes (1, 2, 3, 4) across superpopulations (p=0.4), indicating that this does not explain observed ancestry-associated survival differences. Lastly, AFR patients with schwannomas exhibited significantly worse EFS relative to EUR patients (median EFS 1.4 vs. 1.8 years, HR=11.5, p=0.02; **Figure S4A-B**). However, given the small schwannoma cohort size (N=23) and few number of events (N=9), it is possible that these results may not replicate in larger cohorts.

**Figure 4.**
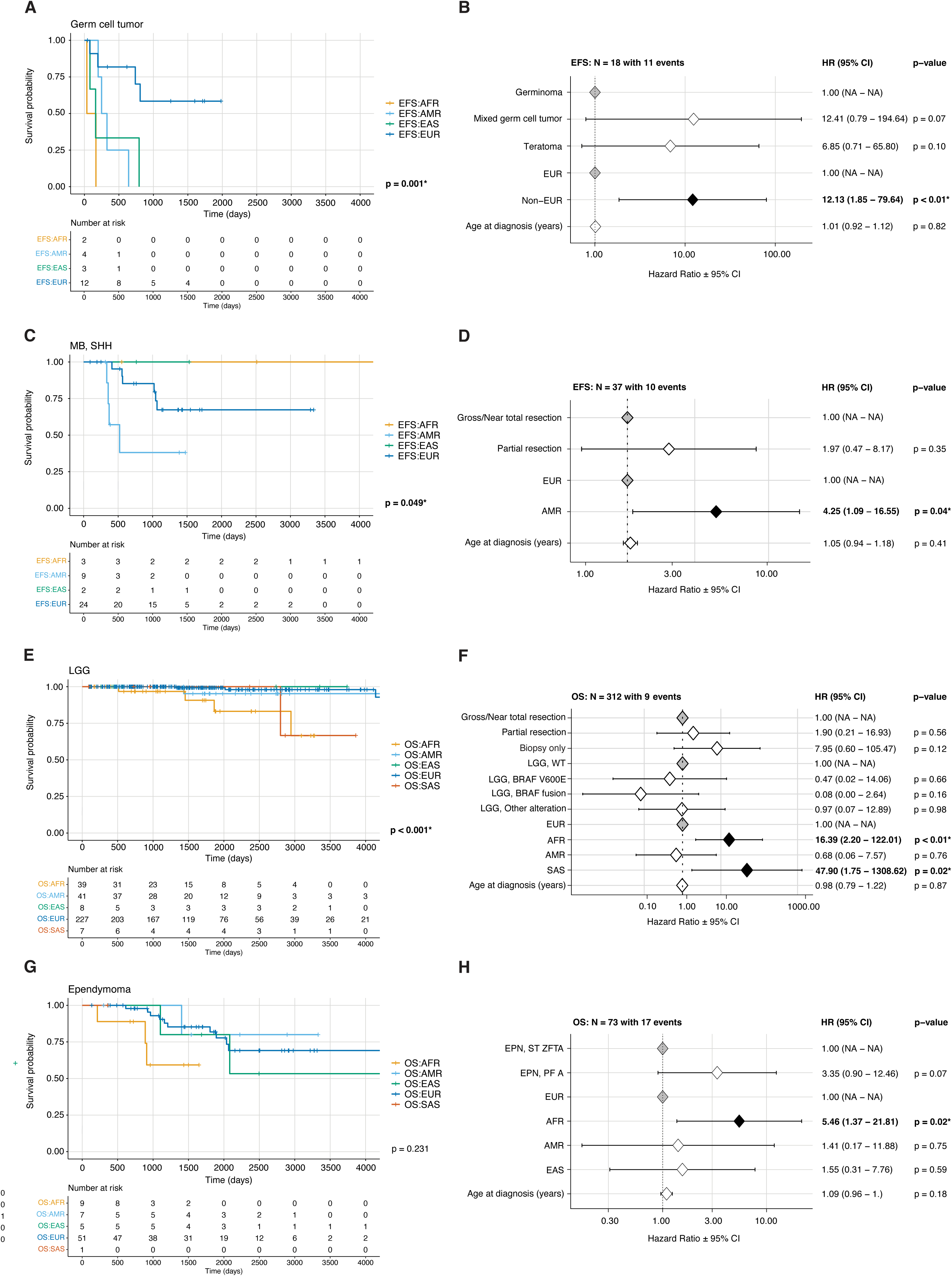
Genetic ancestry-associated overall and event-free survival (EFS) differences in pediatric CNS tumor patients. Kaplan-Meier EFS curve in patients with germ cell (**A**), MB, SHH subtype (**C**), LGG (**E**), and EPN (**G**) tumors by genetic ancestry superpopulation. Cox proportional hazards model forest plots of EFS in patients with germ cell tumors (**B**) or MB, SHH subtype tumors (**D**), including covariates for predicted ancestry, age at diagnosis, and CNS tumor histology (germ cell tumors only). Cox proportional hazards model forest plots of OS in patients with LGG **(F)** and EPN **(H)** tumors, including covariates for molecular subtype, genetic ancestry superpopulation, age at diagnosis, and extent of tumor resection (LGG only). The p-values in the Kaplan-Meier survival curves indicate a significant difference in survival probability across genetic ancestry superpopulations. In the forest plots, grey diamonds represent the reference (control) groups, black diamonds represent a significant difference between a group and the reference (p < 0.05), and white diamonds represent non-significant differences between a group and the reference. Listed for each group are the hazard ratio (HR) with 95% confidence intervals and p-values for each term. A p-value < 0.05 for a given group level in forest plots indicate a significant difference in relative risk of an event relative to that of the reference level of group.

Among patients with LGG, individuals from the AFR superpopulation exhibited significantly worse OS relative to those from the EUR superpopulation (median OS 2.9 vs. 4.2 years, HR=13, p=0.01; **Figure 4C-D**). While we also observed a significantly worse OS in patients from the SAS vs. EUR superpopulation (HR=35, p=0.03), this was driven by a single event among the SAS group, and therefore may not replicate in a larger cohort. When only considering non-*BRAF* altered LGG tumors, the same trend of worse OS in individuals from the AFR superpopulation relative to the EUR superpopulation was observed (HR=49, p=0.01; **Figure S4C&E**). Non-*BRAF* altered tumors were primarily *NF1* or *MAPK*-altered, but we did not find evidence of significant enrichment for any superpopulation when we investigated more specific subtypes, likely due to low numbers per subtype (**Figure S5**). For patients with EPN tumors, we observed significantly worse OS in individuals from the AFR compared to EUR superpopulation (median OS 2.6 vs. 4.7 years, HR=5.5, p=0.02; **Figure 4G-H**), particularly when only considering *ZFTA* fusion-positive EPN (median OS 2.4 vs. 5.1 years, HR=30, p=0.01; **Figure S4D&F**).

We next leveraged PC values derived from somalier ancestry prediction to assess whether these continuous measures of estimated ancestry were associated with patient survival. Among the five derived PC values, we observed significant correlations with estimated probabilities of distinct superpopulations (**Figure S6A**). Most notably, PC1 and PC2 values were significantly positively correlated with estimated AFR and EUR probability, respectively (PC1- AFR: r=0.96, p<1e-10, PC2-EUR: r=0.84, p<1e-10). We ran survival models including PC values as continuous variables and recapitulated several findings observed from survival models that included superpopulation categorical variables (**Figure S6B-C**). For example, increased PC1 values were associated with worse OS in LGG and EPN cohorts and worse EFS in the GCT cohort, while increased PC2 values were associated with better survival in the same cohorts. PC2 values were also significantly associated with worse EFS among patients with ATRT (HR=1.04, p=0.04; **Figure S6D&F**), indicating that EUR ancestry is associated with worse survival. A similar trend of worse EFS in EUR vs. non-EUR patients was observed in categorical superpopulation ATRT EFS models, but this was not statistically significant (non-EUR HR=0.58, p=0.18; **Figure S6E&G**).

### Genetic ancestry superpopulations exhibit distinct treatment patterns

To explore therapeutic approaches employed across genetic ancestry superpopulations, we investigated degree of surgical intervention, clinical trial enrollment, and therapeutic regimens cohort-wide and within tumor histologies. We assessed degree of surgical resection by superpopulation for all major histologies with N > 45 patients, excluding DIPG or DMG (**Figure S7**). Patients from the EAS and SAS superpopulations with mixed neuronal-glial tumors were significantly more likely to undergo total resection relative to other superpopulations of the same histology (EAS: N=2, OR=1.3, FDR=0.01; SAS: N=6, OR=1.3, FDR=0.01; **Figure S7E**).

To determine if this was due to differences in tumor location, we assessed anatomical distributions of tumors by superpopulation (**Figure S8**). Mixed neuronal-glial tumors from EAS and SAS patients were significantly more likely to be in hemispheric brain regions (EAS: N=2, OR=1.2, FDR=0.01; SAS: N=5, OR=1.2, FDR=0.01; **Figure S8E**), which likely contributes to observed more definitive surgical intervention in these populations.

Overall frequency of upfront clinical trial enrollment was significantly associated with genetic ancestry (p=0.001; **Table 1**), with AMR and EUR patients exhibiting significantly higher and lower relative enrollment rates, respectively (AMR: OR=2.1, p=1.7e-04; EUR: OR=0.68, p=0.01; **Figure 5A**). The overall higher rate of AMR superpopulation upfront enrollment was driven in part by patients from DIPG or DMG and EPN tumor cohorts, for which increased rates of AMR superpopulation enrollment were also observed (OR=4.2, p=2.1e-03 and OR=5.3, p=9.4e-03, respectively). To determine if inclusion of PNOC cohort patients was driving observed differences in DIPG or DMG, we reran the analysis excluding this cohort (**Figure S9**). While there was still a trend toward increased upfront enrollment in AMR patients among patients with DIPG or DMG, this was no longer statistically significant (OR=3.7, p=0.06). Genetic ancestry was also significantly associated with rate of upfront enrollment among patients with mesenchymal tumors (p=0.048), with patients from the SAS superpopulation having significantly higher relative rates of enrollment (OR=21.9, p=0.03). And, among patients with HGG, those from the AFR superpopulation were enrolled upfront at significantly higher rates than those from other superpopulations (OR=11.9, p=7.6e-03).

**Figure 5.**
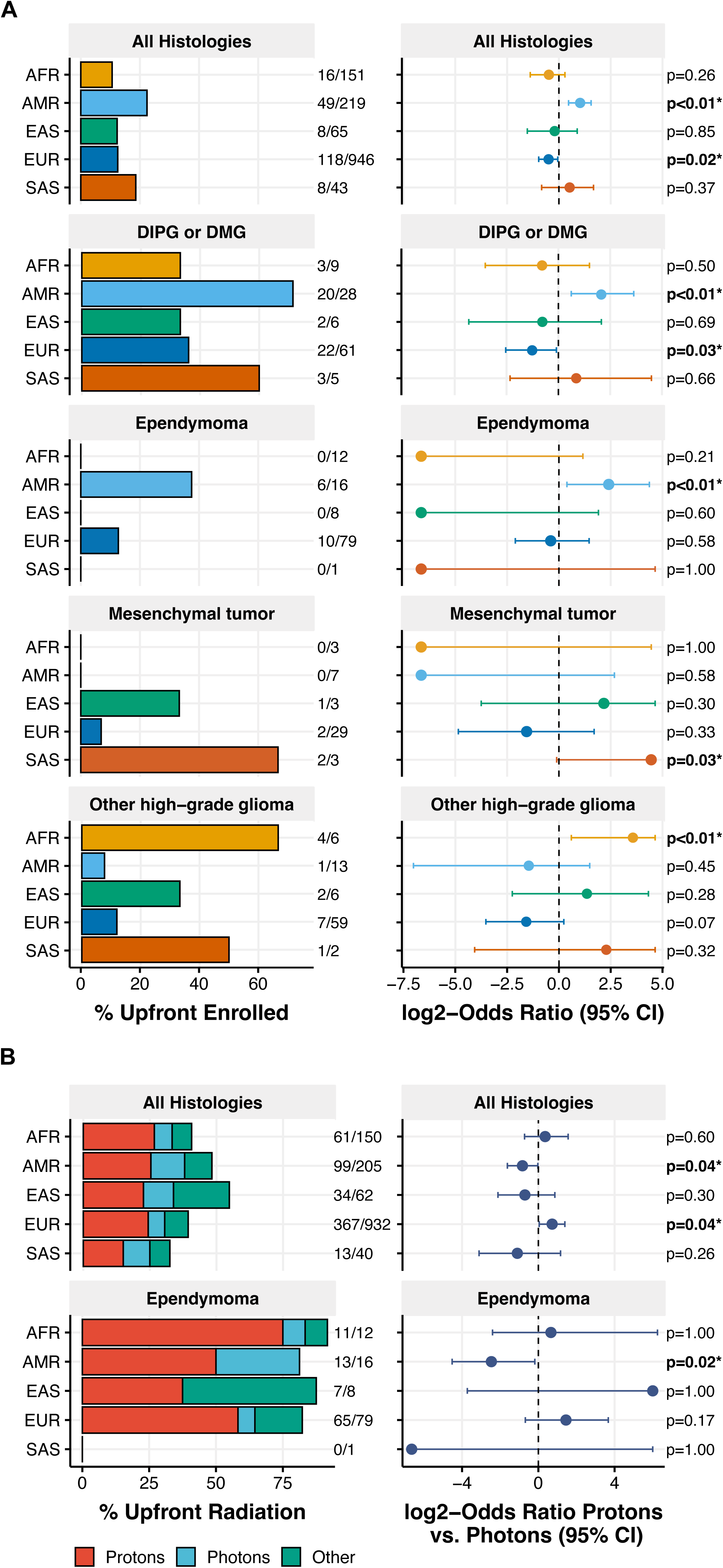
Pediatric CNS tumor treatment frequency by genetic ancestry superpopulation. **A**. Rates of upfront clinical trial enrollment across all tumor histologies and in specific histologies by genetic ancestry superpopulation, and corresponding odds ratio of enrollment in each superpopulation relative to others. **B.** Rates of radiation therapy treatment by radiation type and genetic ancestry superpopulation across all tumor histologies and in patients with EPN. Odds ratios (ORs) indicate likelihood of receiving proton versus photon radiation in superpopulation relative to others, where log_2_-OR > 0 and log_2_-OR < 0 indicate significantly increased likelihood of proton and photon radiation, respectively. Odds ratios and p-values are derived from Fisher’s exact tests.

We further assessed whether frequency of proton versus photon radiation therapy differed across superpopulations. There was a trend toward genetic ancestry-dependent frequency of upfront proton radiation treatment across all tumor histologies (p=0.05). Patients from the EUR superpopulation were significantly more likely to receive proton over photon radiation relative to other superpopulations (OR=1.6, p=0.04; **Figure 5B**) while those of the AMR superpopulation were significantly more likely to receive photon radiation (OR=0.55, p=0.04). Higher rates of photon radiation among the AMR superpopulation were also observed among patients with EPN (OR=0.17, p=0.02).

We incorporated clinical trial and radiation therapy data into survival models to determine if treatment was associated with survival outcomes in tumor histologies for which genetic ancestry was a significant predictor of survival. There was no significant effect of treatment on survival for patients with LGG, EPN, or GCTs, indicating that ancestry-related differences in survival could not be attributed to differences in treatment (**Figure S10A-C**). In patients with SHH-activated MB, radiation type was a significant predictor of EFS (p=0.01), and patients who were treated with photon radiation had higher rates of events relative to those treated with proton radiation (HR=7, p=0.01; **Figure S10D**). However, the survival difference between patients from AMR versus EUR superpopulations was still significant in this model (HR=6.7, p=0.02), indicating that genetic ancestry predicts EFS in this subtype independent of treatment.

## Discussion

In this study, we sought to investigate the influence of genetic ancestry on prevalence of CNS tumor subtypes, treatment access, and survival outcomes in pediatric patients. Our findings revealed significant associations between genetic ancestry superpopulations, prevalence of CNS tumor histologies and molecular subtypes, patient OS and EFS, and upfront treatment approaches. Notably, we also identified superpopulation-specific enrichment of *BRAF* fusion breakpoints among patients with LGG. This work provides a crucial framework for better understanding the contributions of intrinsic germline and tumor genetics and societal components of race and ethnicity in patients with pediatric CNS tumors.

Race and ethnicity are social constructs based on membership in a group sharing cultural and behavioral traits, whereas genetic ancestry is based on variations in genomic structure between groups from similar geographic regions^13,25,26^. The concordance between reported race and ethnicity and predicted genetic ancestry can vary significantly by group^13^. In our work, we observed a significant association between genetic ancestry superpopulation and reported race and ethnicity, consistent with previous findings showing high agreement between reported Black and White race and predicted African and European ancestry, respectively, with larger variation between those of reported American Indian, Alaska Native, or Asian race^13^.

We found significant associations between ancestry superpopulations and cancer group. Patients with ATRT or meningioma tumors were significantly enriched among AFR patients. While previous work has shown an increased incidence of meningiomas among patients of reported Black race^27^, our study is the first to report differences in ATRT incidence by genetic ancestry. Ostrom et al. previously investigated race and reported Hispanic ethnicity in relation to ATRT prevalence, but reported no differences in incidence^28^. We observed significant enrichment of DIPG or DMG tumors among AMR patients, although this was shown to be driven by inclusion of patients from PNOC trials that were disproportionately of AMR ancestry. Additionally, GCTs and schwannomas were significantly enriched in EAS and SAS patients, respectively.

We next report associations between predicted ancestry superpopulation and tumor molecular subtypes. EUR superpopulation patients with LGG diagnoses had tumors enriched with non-*BRAF* altered subtypes associated with higher recurrence rates and worse progression-free survival^21^. EAS superpopulation patients with HGG were more likely to harbor *TP53* mutations relative to other superpopulations. While this finding has not been reported previously in pediatric CNS tumors, a breast cancer study reported higher rates of *TP53* mutations in Asian patients compared to Caucasian patients with ER+ tumors^29^.

In our analysis of LGG breakpoint type frequency, we found that 93.6% of patients harbored one of the four common *KIAA1549::BRAF* breakpoint types^22^. EAS and SAS patients had tumors significantly enriched for the common 15:09 breakpoint, while AMR patients had tumors enriched for rare fusion breakpoints. Previous work has shown that rare breakpoints are associated with worse clinical outcomes in patients with LGG^22^. We recapitulate and extend these findings, showing that rare *KIAA1549::BRAF* breakpoints tumors are enriched in anatomic locations that are inherently more difficult to resect (suprasellar and optic pathway). Interestingly, we also observed enrichment of tumors with the common 15:09 breakpoint in regions less likely to be resected (spinal and suprasellar regions); however, 15:09 breakpoint tumors were not associated with worse EFS relative to other common breakpoint tumors. Lastly, both 15:09 and rare breakpoints were significantly more likely to be diagnosed as PMAs, which have been shown to exhibit worse progression-free and overall survival relative to PAs^30^.

In this cohort, we found that patients of predicted non-European ancestry exhibited significantly worse OS and EFS compared to patients of predicted European ancestry in certain tumor histologies and molecular subtypes. Patients of non-EUR ancestry with GCT and of AMR ancestry with SHH medulloblastomas had worse EFS compared EUR patients with the same diagnoses. AFR superpopulation patients with LGG or EPN showed significantly worse OS compared to EUR patients, consistent with prior findings of lower survival among Black children with these tumors^31,32^. Furthermore, these observations remained when restricting analyses to specific molecular subtypes (non-*BRAF* altered LGG and *ZFTA* fusion-positive EPN, respectively). When incorporating ancestry prediction PC values into survival models as an alternative strategy to assess ancestry-associated survival trends, we recapitulated all significant findings from survival models with superpopulation categorical variables. However, we also observed a significant association between PC2—associated with increased EUR ancestry probability—and worse EFS in patients with ATRT. ATRT EFS models that included categorical population variables did not reveal a significant association between the EUR ancestry and EFS, emphasizing the importance of measuring genetic ancestry using alternative strategies beyond broad superpopulation assignment. To our knowledge, this is the first report of worse outcomes among EUR patients with ATRT compared to non-European ancestries.

Our study also found varying upfront clinical trial enrollment frequencies by genetic ancestry superpopulation. This is encouraging, as it highlights the importance of including patients from diverse genetic backgrounds in clinical research and may represent contemporary changes that better facilitate equitable enrollment in trials and research activities^33,34^. In contrast, EUR superpopulation patients were significantly more likely to receive proton over photon radiation, while the converse was observed among AMR patients. This result aligns with a previous study assessing frequency of proton vs. photon radiation therapy in patients with pediatric tumors, which reported higher incidence of proton radiation therapy among non-Hispanic White patients relative to historically marginalized race groups^6^. While radiation type was not consistently associated with survival differences in our cohort, photon radiotherapy has been associated with higher risk of adverse side effects relative to proton radiotherapy^35^. This work emphasizes the need for assessing frequency of other events that may arise due to disparities in treatment.

Although our work included patients from a diverse set of genetic and predicted ancestral backgrounds, our data came from patients within the United States. Thus, our cohort was significantly enriched for the European superpopulation, with limited numbers of historically marginalized racial and ethnic groups in our study (i.e., AI/AN and NHPI). Expanding access to international pediatric datasets with potentially unique SDoH considerations will be critical for including diverse representation in future research.

While the inclusion of molecular subtypes in our analyses allowed for a more nuanced exploration of tumor risk across superpopulations, our sample sizes for several subtypes were limited. We report our findings with full consideration of these small sample sizes, which we hope to mitigate with ongoing enrollment and larger sample sizes in planned follow-up reports. Further, the discovery of novel molecular features and further subdivision of current molecular subtypes may lead to reduced power to detect ancestry associated prevalence and outcomes. For example, the *SHH* subgroup of medulloblastoma is now categorized into four subtypes based on molecular features and are associated with different survival outcomes^36^. In addition, pathogenic germline variants are known to play a role in the development of CNS tumors and may be enriched in certain genetic superpopulations^34,37^. Epigenetic processes are also known to contribute to cancer development, and differential DNA methylation patterns among racial groups at birth, notably in cancer pathway genes, has previously been reported^38^. However, we did not explore the contribution of germline findings and DNA methylation patterns to observed superpopulation differences in this study. Our data too may be affected by sample bias from locoregional population enrichment in catchment areas of our CBTN/PNOC enrolling sites. For instance, PNOC sites demonstrated higher correlation of patients from AMR ancestry with DMG/DIPG; however, this may be more reflective of patient concentration from enrolling sites in California, which is known to include high populations of Indigenous populations and Mexican populations that may be reflective of AMR ancestry^39,40^. Such enrichment may bias correlation, as was demonstrated when repeat analyses were done with removal of the PNOC cohort. It is also important to consider distinct components of genetic ancestry within Admixed and Hispanic populations across different regions of the US and how this may affect incidence and outcome of brain tumors at the locoregional level, as has been recently reported^41^. As more data become available through CBTN, PNOC, and other consortia, we expect to expand our analyses to include additional patients, integrate germline and somatic variation, and further assess genetic subpopulations.

Importantly, our work did not consider the contributions of SDoH to survival, such as socioeconomic status, insurance status, and time to diagnosis, which are well-known to impact survival^2,42,43^. Although information on these key SDoH was not available for our retrospective analysis, the CBTN and PNOC aim to include these factors, such as the childhood opportunity index or area deprivation index in future prospective and longitudinal data collection. Historically, there has been a lack of reporting of race and ethnicity in clinical trials and we strive to improve this in the pediatric cancer field, as it is essential to understanding clinically-relevant associations with race and/or ethnicity. Future work should explore these societal factors alongside genetic contributions to cancer incidence and survival, as well as access to clinical trials and treatment regimens. While we used reported race and ethnicity retrospectively collected from electronic medical record abstraction, prospective data collection of demographic information using a patient’s self-report may more accurately reflect patients’ identities.

Our work revealed several new findings in pediatric CNS tumor histologies and molecular subtypes across genetic superpopulations, highlighting associations with survival and upfront treatment approaches. To further improve equity in care and outcomes for children with CNS malignancies, additional research is needed to delineate the extent to which race and ethnicity differences are driven by societal determinants or tumor biology and molecular subtypes related to genetic ancestry.

## Data Availability

Raw sequencing data can be accessed at dbGaP accession number phs002517 or data access request to CBTN. All data and code used to perform analyses and generate figures for this manuscript can be found at https://github.com/rokitalab/pbta-genetic-ancestry.

## Acknowledgements and Funding

We have deep gratitude to the patients and families who contributed tissue and data through participation in studies by the CBTN, PNOC, and other consortia such as Oligo Nation. C.K. is the recipient of the Robert A. Winn Diversity in Clinical Trials Career Development Award, established by Bristol Myers Squibb Foundation. This work was also supported in part by the CBTN, the Chad Tough Foundation, and the Division of Neurosurgery at Children’s Hospital of Philadelphia.

## Author Contributions

Conceptualization: CK, JLR, RJC, SMW

Methodology: RJC, JLR, MAB, CK

Software: MAB, RJC, CB

Validation: RJC, JLR, ZG, AC, ASN

Formal Analysis: BZ, CZ, RJC, JLR

Investigation: RJC, JLR, CK

Resources: CK, JLR, PBS, ACR, SMu, MP

Data Curation: RJC, JLR, CK, SMc, ED, NVK, AP

Writing – Original Draft: RJC, CCG, CK, JLR

Writing - Review & Editing: RJC, CCG, CK, JLR, SMc, TBD, AC, ED, AJW, AP, MAB, SMW

Visualization: RJC, CCG, JLR, CK Supervision: JLR, CK

Project Administration: JLR, CK Funding Acquisition: PBS, ACR, CK

## Data and Code Availability

Raw data for the PBTA and PNOC can be accessed at dbGaP accession number phs002517 or data access request to cbtn.org and pnoc.us. All data and code used to perform analyses and generate figures for this manuscript can be found at https://github.com/rokitalab/pbta-genetic-ancestry.

