## Supplemental Legends for "Genetic ancestry superpopulations show distinct prevalence and outcomes across pediatric central nervous system tumors from the PBTA and PNOC"

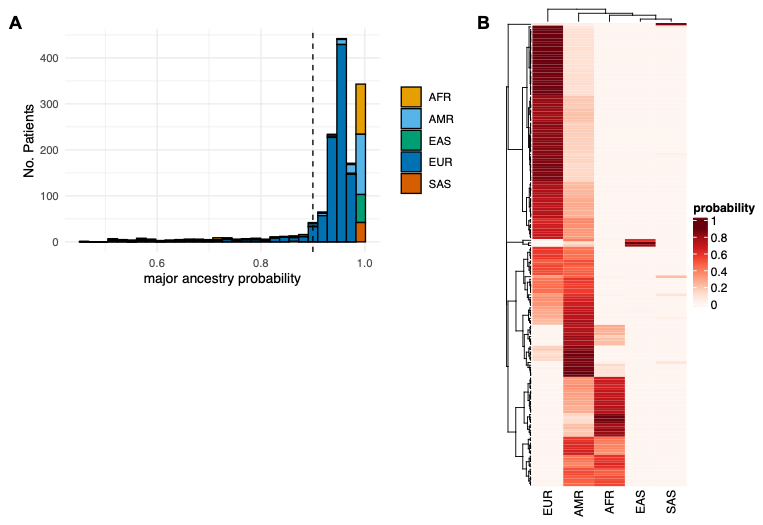

**Figure S1.** (**A**) Histogram of major genetic ancestry superpopulation probability in pediatric CNS tumor cohort. Vertical dashed line indicates probability = 0.9. (**B**) Heatmap of genetic ancestry superpopulation probabilities in patients with major predicted genetic ancestry probability <0.9 (N = 178). Clustering was performed using the “complete” agglomeration method and Euclidean distance measure.

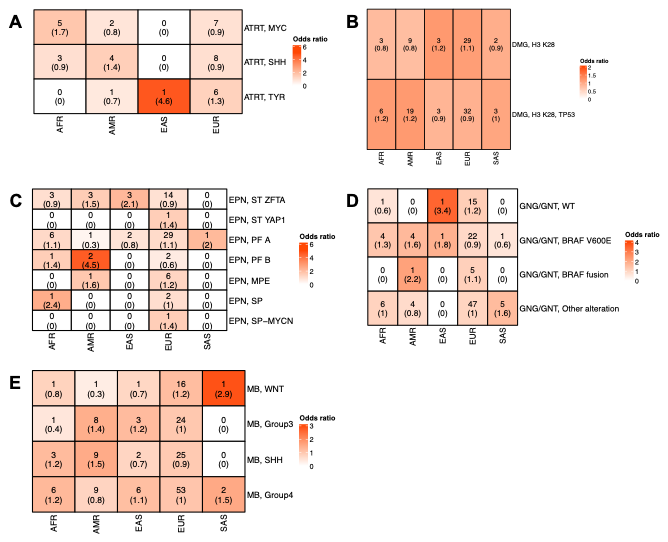

**Figure S2.** **A-E.** Count of genetic ancestry superpopulations within Atypical Teratoid Rhabdoid Tumor (ATRT, **A**), Diffuse midline glioma (**B**), ependymoma (**C**), mixed neuronal-glial tumor (**D**), and medulloblastoma (**E**) molecular subtype cohorts, with Fisher’s exact test-derived odds ratios in parentheses. Box colors are weighted by odds ratio. Stars denote FDR < 0.05.

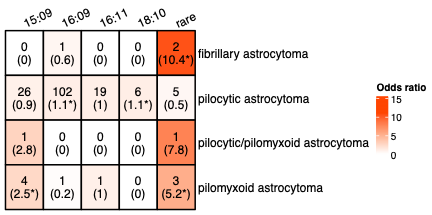

**Figure S3.** Count of *KIAA1549::BRAF* fusion breakpoint group tumors among low-grade glioma diagnoses, with Fisher’s exact test-derived odds ratios in parentheses. Box colors are weighted by odds ratio. Stars denote FDR < 0.05.

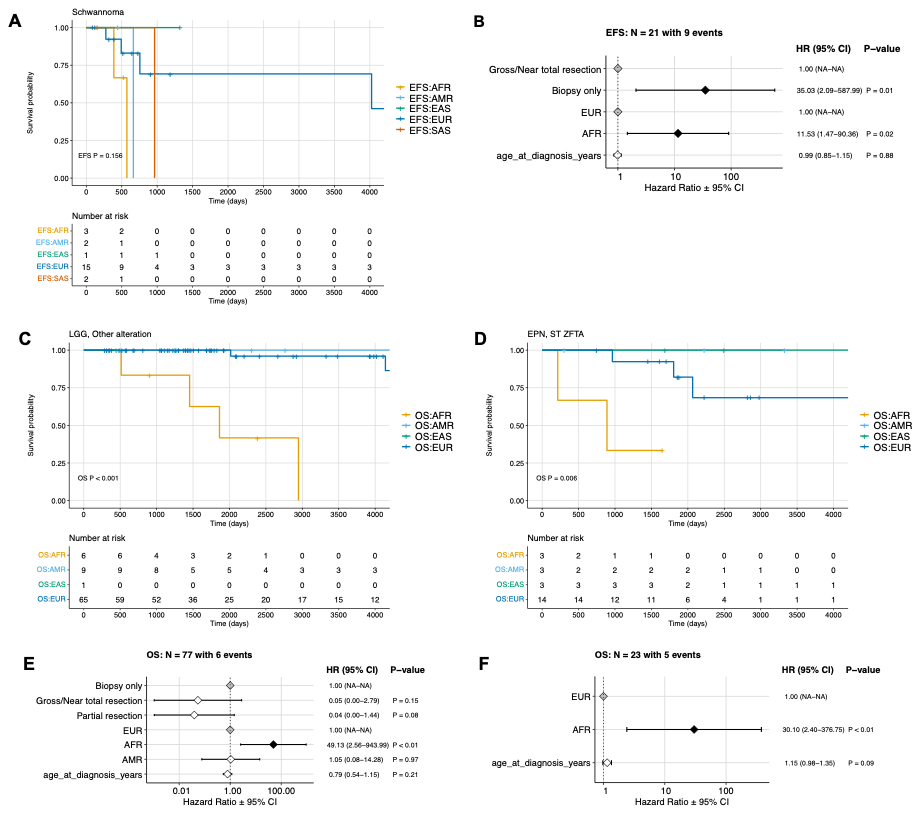

**Figure S4. Genetic ancestry-associated survival differences in pediatric CNS tumor patients. A**. Kaplan Meier event-free survival (EFS) curve in schwannoma cohort. **B.** Cox proportional hazards model forest plots of EFS in schwannoma cohort, including covariates for extent of tumor resection, age at diagnosis, and genetic ancestry superpopulation. **C-D.** Kaplan Meier overall survival (OS) curve in (**C**) LGG, non-*BRAF* altered subtype cohort and (**D**) EPN, *ZFTA* fusion-positive subtype cohort. **E-F.** Cox proportional hazards model forest plots of overall survival in (**E**) LGG, non-*BRAF* altered subtype and (**F**) EPN, *ZFTA* fusion-positive patients, including covariates for predicted ancestry, age at diagnosis, and extent of tumor resection (LGG only). P < 0.05 in Kaplan-Meier survival curves indicates a significant difference in survival probability between genetic ancestry superpopulations. A p-value < 0.05 for a given group level in forest plots indicates a significant difference in relative risk of an event relative to that of the reference level of group.

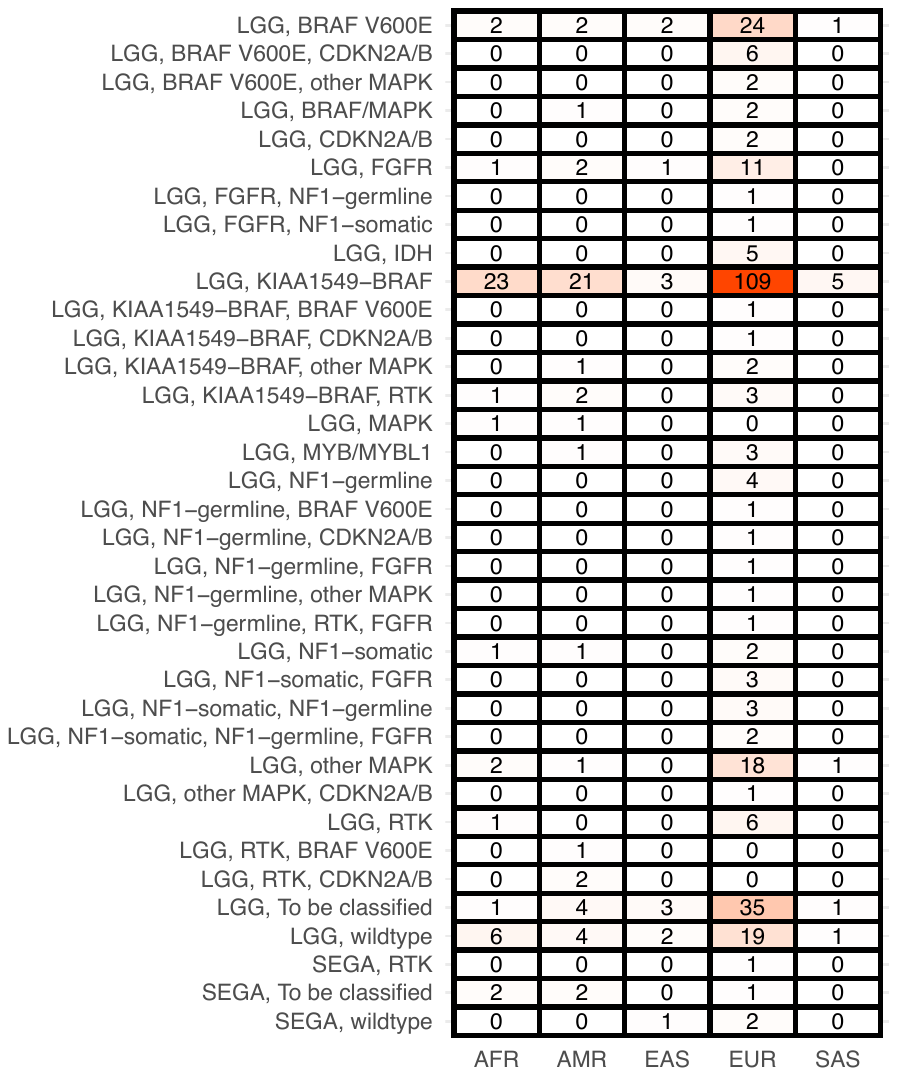

**Figure S5.** Distribution of LGG molecular subtypes by genetic ancestry superpopulation. Box colors are weighted by N.

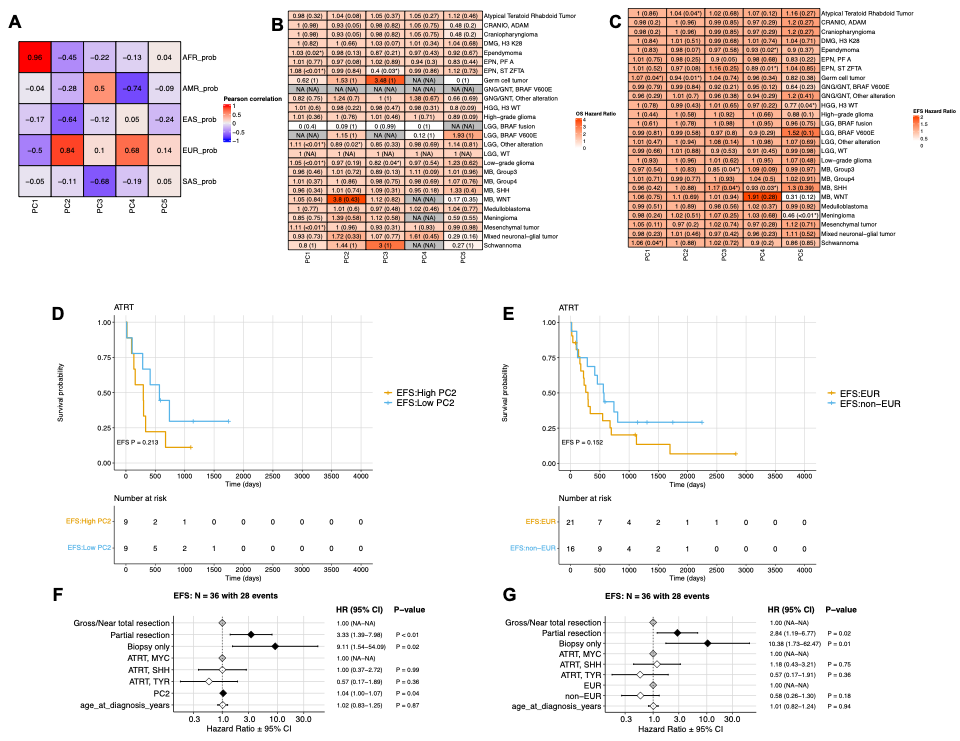

**Figure S6.** **A.** Heatmap of Pearson correlation coefficients between somalier-derived principal component (PC) values and probability of genetic ancestry superpopulations. **B-C.** Heatmaps of overall survival (OS, **B**) and event-free survival (EFS, **C**) hazard ratios of PC value covariates derived from cox proportional hazards survival models, with corresponding p-values in parentheses. Stars denote p<0.05. **D-E.**  Kaplan-Meier EFS curve in ATRT cohort, with samples stratified by **D)** high (upper quartile) and low (lower quartile) PC2 values, and **E)** EUR and non-EUR superpopulation assignment. F-G. Cox proportional hazards model forest plots of EFS in ATRT cohort, including covariates for **F)** PC2 value and **G)** EUR superpopulation assignment (EUR or non-EUR). A p-value < 0.05 for PC2 value indicates a significant association between increases in PC2 value and risk of event. A p-value <0.05 for a given categorical group level in forest plots indicates a significant difference in relative risk of an event relative to that of the reference level of group.

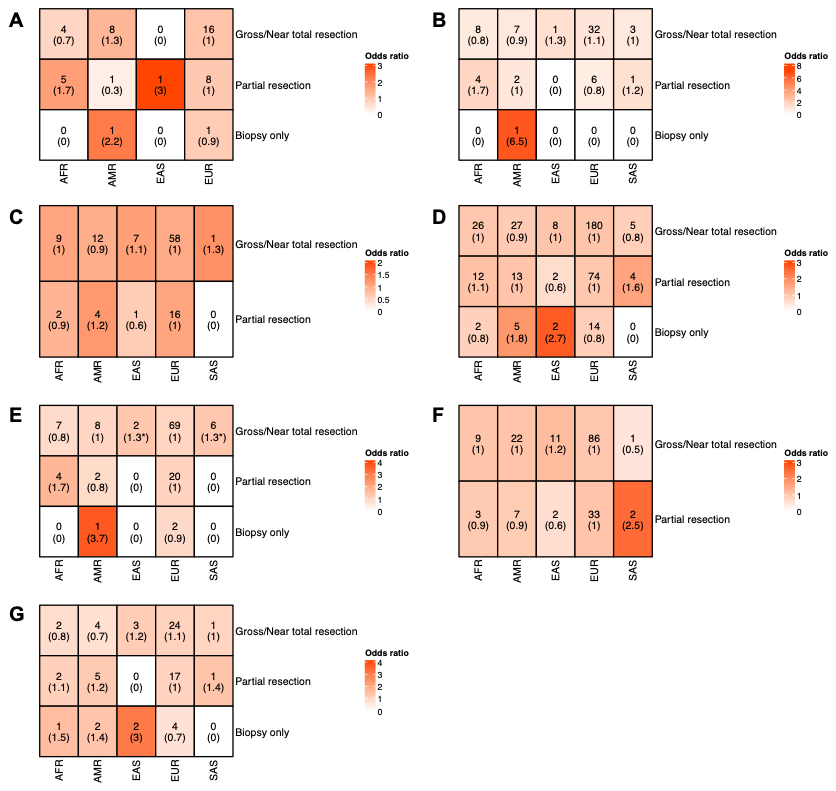

**Figure S7.** Count of genetic ancestry superpopulation members among surgical resection groups, with Fisher’s exact test-derived odds ratios in parentheses, in the following tumor histologies: Atypical Teratoid Rhabdoid tumor (**A**), craniopharyngioma (**B**), ependymoma (**C**), low-grade glioma (**D**), mixed neuronal-glial tumors (**E**), medulloblastoma (**F**), and high-grade glioma (**G**). Box colors are weighted by odds ratio. Stars denote FDR < 0.05.

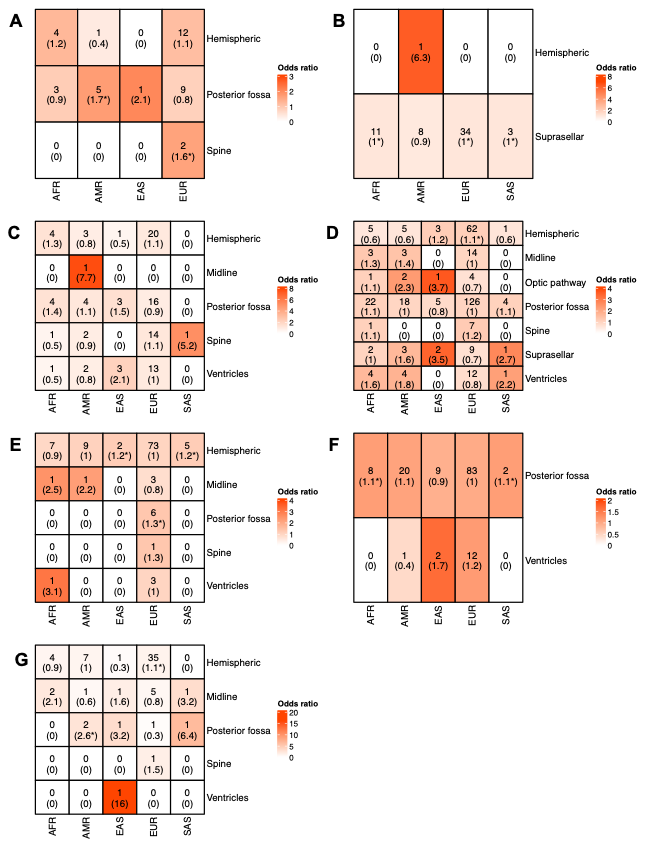

**Figure S8.** **A-E.** Count of genetic ancestry superpopulation members by anatomical region for the following tumor histologies: Atypical Teratoid Rhabdoid tumor (**A**), craniopharyngioma (**B**), ependymoma (**C**), low-grade glioma (**D**) mixed glial-neuronal tumors (**E**), medulloblastoma (**F**), and high-grade glioma (**G**). Box colors are weighted by odds ratio. Stars denote FDR < 0.05.

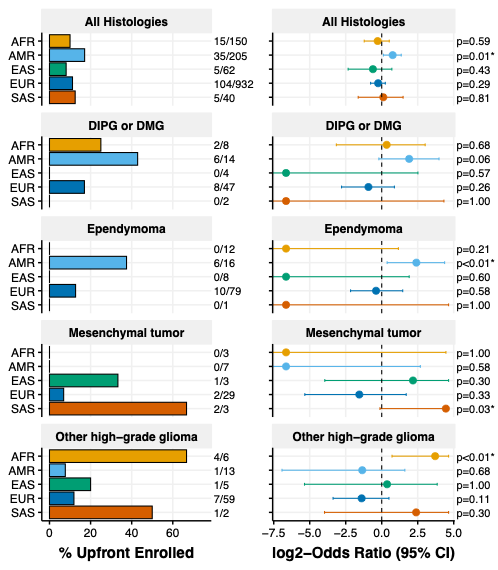

**Figure S9.** Rates of upfront clinical trial enrollment by genetic ancestry superpopulation when PNOC clinical trial patients are excluded, and corresponding odds ratio of enrollment in each superpopulation relative to others. Only histologies for which significant enrichment was observed are displayed. Odds ratios and p-values are derived from Fisher’s exact tests.

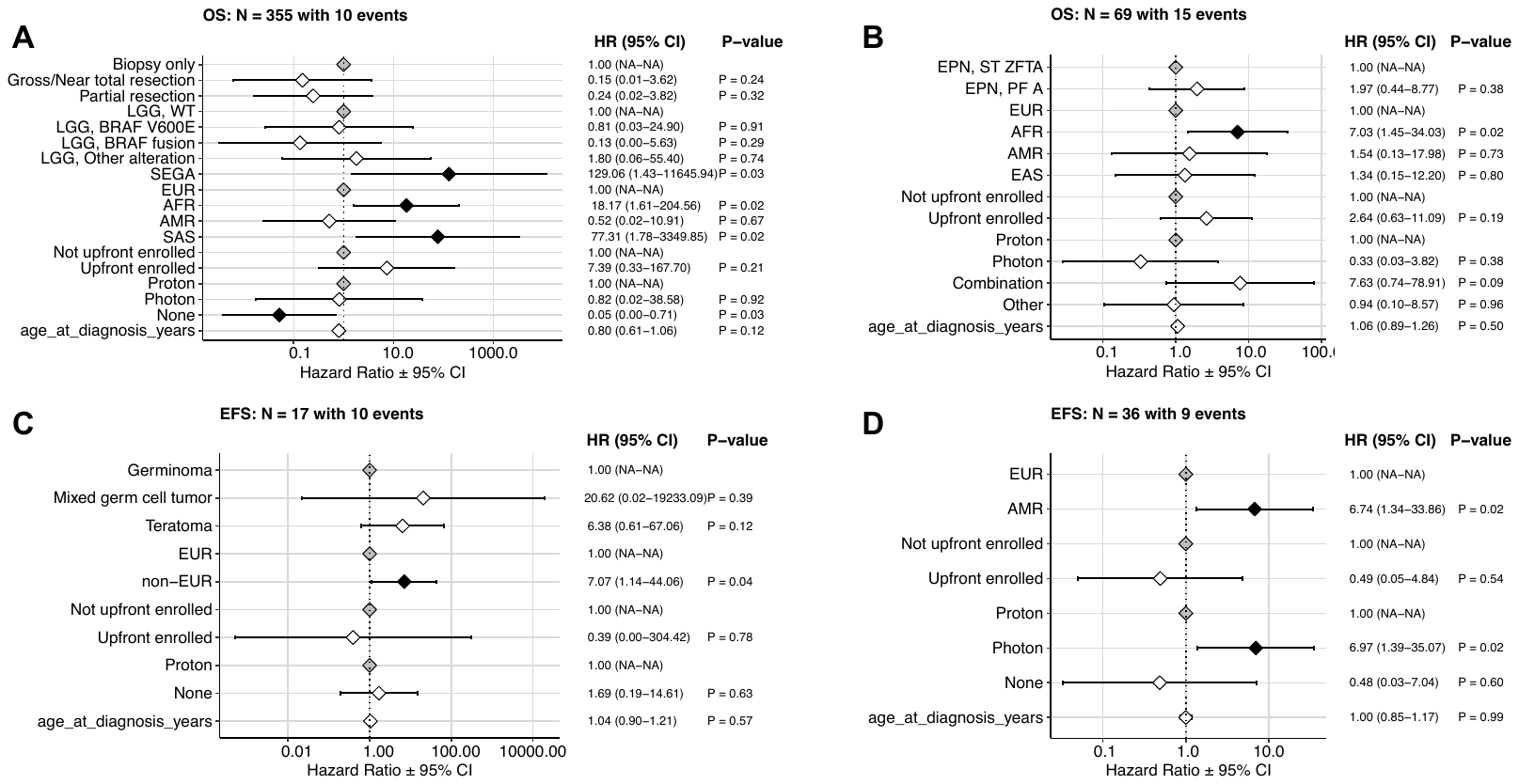

**Figure S10.** Cox proportional hazards model forest plots of overall survival (OS) and event-free survival (EFS) in patients with LGG (**A**), EPN (**B**), germ cell tumors (**C**), and medulloblastoma SHH subtype (**D**). Covariates include predicted ancestry, upfront clinical trial enrollment, radiation type, extent of tumor resection (LGG only), molecular subgroup (LGG, ependymoma), and cancer group (germ cell tumors).
